# Glucagon-like peptide-1 receptor agonists (GLP-1RA) for neuroprotection following aneurysmal subarachnoid haemorrhage (aSAH): a scoping review

**DOI:** 10.1101/2025.07.23.25332036

**Authors:** Matt Thomas, Svetlana Mastitskaya, Sam Parker, Ruth Cookson, Lucy Holmes, Aidan Marsh, Aravind Ramesh, Sarah Rudd, Mario Teo, Alex Mortimer

## Abstract

**Objective:** The objective of this scoping review is to understand the extent and type of evidence in relation to the use of glucagon-like peptide-1 receptor agonists (GLP-1RA) for neuroprotection in aneurysmal subarachnoid haemorrhage (aSAH).

**Introduction:** The individual and societal costs of aSAH remain high. Effective neuroprotection would reduce morbidity and mortality but there are uncertainties around both established and emerging therapies. GLP-1RA show promise as neuroprotective drugs in other forms of acute and chronic brain injury and could be repurposed for aSAH.

**Inclusion criteria:** Animal and human studies of the use of GLP-1RA for aSAH will be included. Key exclusions are traumatic or non-aneurysmal subarachnoid haemorrhage, or the use of multi-agonist drugs.

**Methods:** Searches were conducted in Embase (Ovid), Medline (Ovid), Cochrane Central Register of Controlled Trials (Wiley) and the World Health Organisation’s International Clinical Trials Registry Platform on 13th June 2025 with no limits applied. Screening and data extraction was performed by two independent reviewers.

**Results:** After de-duplication 593 records were screened, 50 selected for full text review and 5 included in this review. GLP-1R were shown to be highly expressed in neurones and microvascular endothelial cells after aSAH. Administration of GLP-1RAs to rats affected by aSAH improved functional recovery. Furthermore, aSAH was reported to increase cerebral hemisphere oedema, blood-brain barrier permeability, cell death and inflammation, all of which were reversed by GLP-1RA treatment. Murine studies highlight potential mechanisms for these beneficial effects including inhibition of ferroptosis, downregulation of apoptosis, and upregulation of SIRT1 pathways. A human observational studies shows a correlation between higher SIRT1 levels and better neurological outcomes.

**Conclusion:** The limited available evidence suggests a potential neuroprotective role for GLP-1RAs after aSAH. There is a need for extensive further research to determine the efficacy and safety of GLP1-RAs for neuroprotection in aSAH.

## Introduction

Stroke is the third most common cause of death globally, and the fourth most common cause of disability adjusted life years. Aneurysmal subarachnoid haemorrhage (aSAH) is the third most common form of stroke, accounting for around 5% of the total, with 0.7 million (95% uncertainty interval 0.5 – 0.8 million) people affected annually worldwide. Overall incidence is 8.3 (7.3 – 9.5) cases per 100,000 population and mortality 4.2 (3.7 – 4.8) per 100,000 with considerable geographic variation.^1^

Aneurysmal subarachnoid haemorrhage has a younger peak incidence and increasing prevalence in people under 70 years old.^1^ Most survivors have some neurological impairment; only one-third of those in work at the time of ictus can return fully at 2-4 years. ^2,3^ Although the high case fatality rate appears to be decreasing over time,^4,5^ this likely implies increasing demand for resources for rehabilitation and a greater societal cost of disability with the loss of economically productive individuals. For example, the annual cost to the United Kingdom was estimated at £510 million in 2010.^6^ This makes it essential to find effective treatments to improve outcome from aSAH.

Neuroprotection is the preservation, salvage or recovery of central nervous system tissue after acute insult; effective neuroprotective treatments will improve outcomes by reducing death and disability. The current standard neuroprotective treatment for aSAH, nimodipine, is recommended by recent guidelines.^7,8^ The assessment of the quality of evidence and the weights of the recommendations differ between guidelines but there is agreement on the limitations arising from the age of the data and the uncertainties around key outcomes.

There are also practical concerns with nimodipine, including hypotension, vasoplegia, a short half-life and pharmacokinetic variability, that has led to extensive investigation of alternatives.^9^

As yet, none of these alternatives are in widespread use in clinical practice and there remains a large unmet need to find effective agents to improve outcomes.^10^ Drug candidates investigated to date show a lack of effect on long term functional outcomes.^11–13^ This may reflect a focus on a single relatively late component (radiologically documented arterial narrowing or “vasospasm”) that oversimplifies the multiple complex pathophysiological processes set in motion at the time of ictus.^14,15^ The opportunity remains for new classes of potentially neuroprotective drugs to be investigated in aSAH that target processes beyond vasospasm.

Glucagon-like peptide-1 receptor agonists (GLP-1RA) are such a class of drug. GLP-1 is an incretin hormone produced by the pancreas, gut and central nervous system with its receptor (GLP-1R) widely distributed in most organ systems.^16^ Although developed as a treatment for type 2 diabetes to harness its pancreatic effects, GLP-1RA have since been shown to have a wide range of effects throughout the body.^17^ Actions of GLP-1RA that may be relevant to the pathophysiology of aSAH include modulation of endothelial function, mitigation of ischaemia-reperfusion injury, anti-inflammatory effects, regulation of blood glucose and inhibition of apoptosis, as well as promotion of synaptic plasticity and axonal regeneration.^18,19^ All are potentially neuroprotective and there is some evidence of benefit in animals.^20^ In addition they are licensed for other indications in humans, with extensive use demonstrating a low incidence of serious adverse events. Attention to date has largely focused on the use of GLP-1RA for neuroprotection in acute ischaemic stroke and neurodegenerative conditions^21–23^ and whether repurposing for use in aSAH would translate into clinical benefit is as yet unknown.

A preliminary search of PROSPERO, the Cochrane Database of Systematic Reviews, the Open Science Framework and JBI Evidence Synthesis was conducted and no current or underway systematic reviews or scoping reviews on the topic were identified. Based on this search, the fact that GLP-1RAs have mainly been examined in other forms of neurological injury and the observation that current reviews of emerging treatments for aSAH do not include GLP-1RA ^10,24–26^ a scoping rather than systematic review was deemed an appropriate choice.^27^ The aims of this review are to assess the extent of pre-clinical and clinical literature regarding the use of GLP-1RA for neuroprotection in aSAH, identify key knowledge gaps, and serve as a precursor for future reviews and clinical studies including randomised controlled trials.

## Methods

The scoping review has been conducted in accordance with the JBI methodology for scoping reviews.^28^ The review was registered prospectively on 4th October 2024 on the Open Science Framework (osf.io/v95d7).

### Review Question

The review question is: “What is the potential role of GLP-1RA in the management of aneurysmal subarachnoid haemorrhage?”

### Inclusion criteria

#### Participants

Human and animal studies will be included. All study designs, comparators (including no comparators), and study outcomes will be considered.

#### Concept

Studies of aneurysmal subarachnoid haemorrhage will be included. Non-aneurysmal subarachnoid haemorrhage, including traumatic and perimesencephalic subarachnoid haemorrhage, will be excluded.

Glucagon-like peptide-1 receptor agonists, licensed, unlicensed and experimental will be considered. Multi-agonists will be excluded.

#### Context

Aneurysmal subarachnoid haemorrhage is primarily a disease of adults and is often managed in Intensive Care Units; these will not however be restrictions applied to this review. There will be no restrictions based on culture, location or any protected characteristic.

#### Types of sources

This scoping review considers both experimental and quasi-experimental study designs in humans and animals including randomized controlled trials, non-randomized controlled trials, before and after studies and interrupted time-series studies. In addition, analytical observational studies including prospective and retrospective cohort studies, case-control studies and analytical cross-sectional studies will be suitable for inclusion. This review will also include relevant descriptive observational study designs including case series, individual case reports and descriptive cross-sectional studies.

The reference lists of all articles retrieved for full text analysis as well as the authors’ personal collections will be searched for additional information sources.

#### Search strategy

A systematic search of four electronic bibliographic databases (Embase (Ovid), Medline (Ovid), Cochrane Central Register of Controlled Trials (Wiley) and the World Health Organisation’s International Clinical Trials Registry Platform (ICTRP)) was created in conjunction with a medical librarian. The search was conducted on the 13th June 2025 with no limits or filters applied, in line with Cochrane Handbook ^29^ and JBI Manual for Evidence Synthesis^28^ recommendations and other systematic reviews investigating the use of GLP-1 receptor agonists.^30,31^ ^32^ The search strategy includes keyword and subject heading search terms relating to subarachnoid haemorrhage and GLP-1RA’s (including exenatide or liraglutide or albiglutide or dulagutide or lixisenatide or semaglutide or tirzepatide; see Appendix I).

### Study/Source of evidence selection

Following the search, all identified citations were collated and uploaded into Rayyan (https://www.rayyan.ai/) and duplicates removed. Titles and abstracts were screened by two independent reviewers with discrepancies adjudicated by a third reviewer. The full text of selected citations were assessed in detail against the inclusion criteria by two independent reviewers with reasons for exclusion recorded and disagreements resolved by a third reviewer.

### Data extraction

Data was extracted from papers included in the scoping review by two independent reviewers using a data extraction tool developed by the reviewers from the JBI data charting template (see Appendix II).^28^ Data extraction was checked and discrepancies resolved by a third reviewer.

### Data analysis and presentation

Evidence is presented as narrative summaries categorised according to species (human/animal) and outcome and has not been critically appraised or subject to statistical analysis (including meta-analysis).

## Results

### Study inclusion

A total of 887 records were identified by the search. After de-duplication 593 titles and abstracts were screened for inclusion with 543 excluded. Fifty records were obtained in full text with 45 subsequently excluded and 5 studies included in this review. The PRISMA flow diagram with reasons for exclusion is shown in Figure 1.

**Figure 1:**
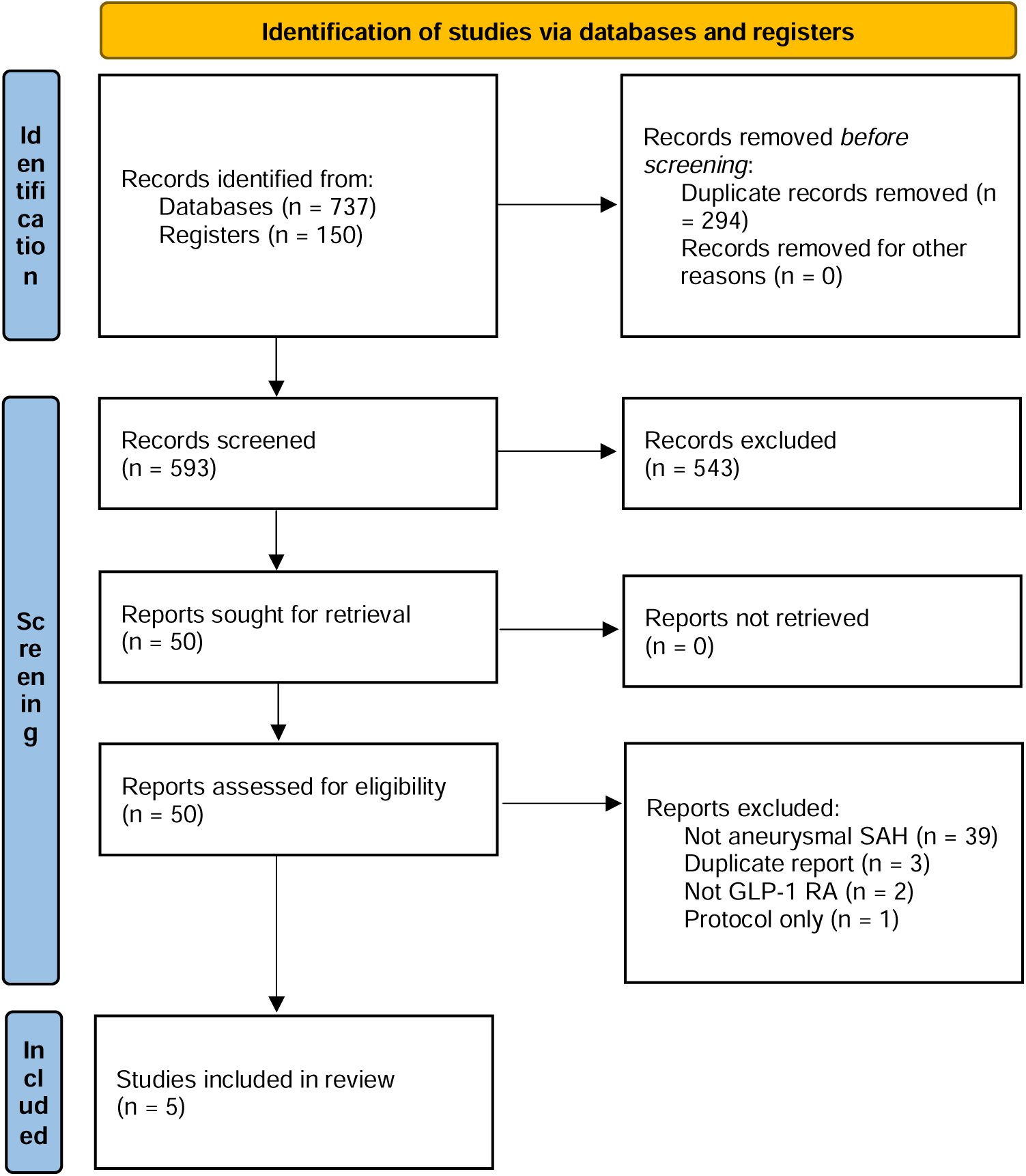
PRISMA flow diagram Automation tools were not used. GLP-1 RA: glucagon-like peptide 1 receptor agonist; SAH: subarachnoid haemorrhage Source: Page MJ, et al. BMJ 2021;372:n71. doi: 10.1136/bmj.n71. This work is licensed under CC BY 4.0. To view a copy of this license, visit https://creativecommons.org/licenses/by/4.0/

### Characteristics of included studies

Five studies were included in this scoping review.^20,33–36^ Three were animal studies (in rats),^33–35^ one reported findings from both animal (mice) and human investigations,^20^ and one was a human trial.^36^ All animal studies were interventional, using the endovascular perforation model of aSAH.^20,33–35^ One human study was an observational case-control study^20^ and one an interventional cohort study;^36^ both were single centre and prospective.

Two animal studies used exendin-4,^33,34^ one liraglutide^35^ and one semaglutide.^20^ The interventional human study used exenatide (a synthetic analogue of exendin-4).^36^ Outcomes common to the animal studies include neurobehavioural function, cerebral oedema and investigation of potential mechanisms of effect.^20,33–35^ Outcomes in the interventional human study predominantly relate to glucose control;^36^ the observational human study examines the association of a potential biomarker (sirtuin-1) with six month functional outcome.^20^

Key characteristics are given in Table 1.

**Table 1:**
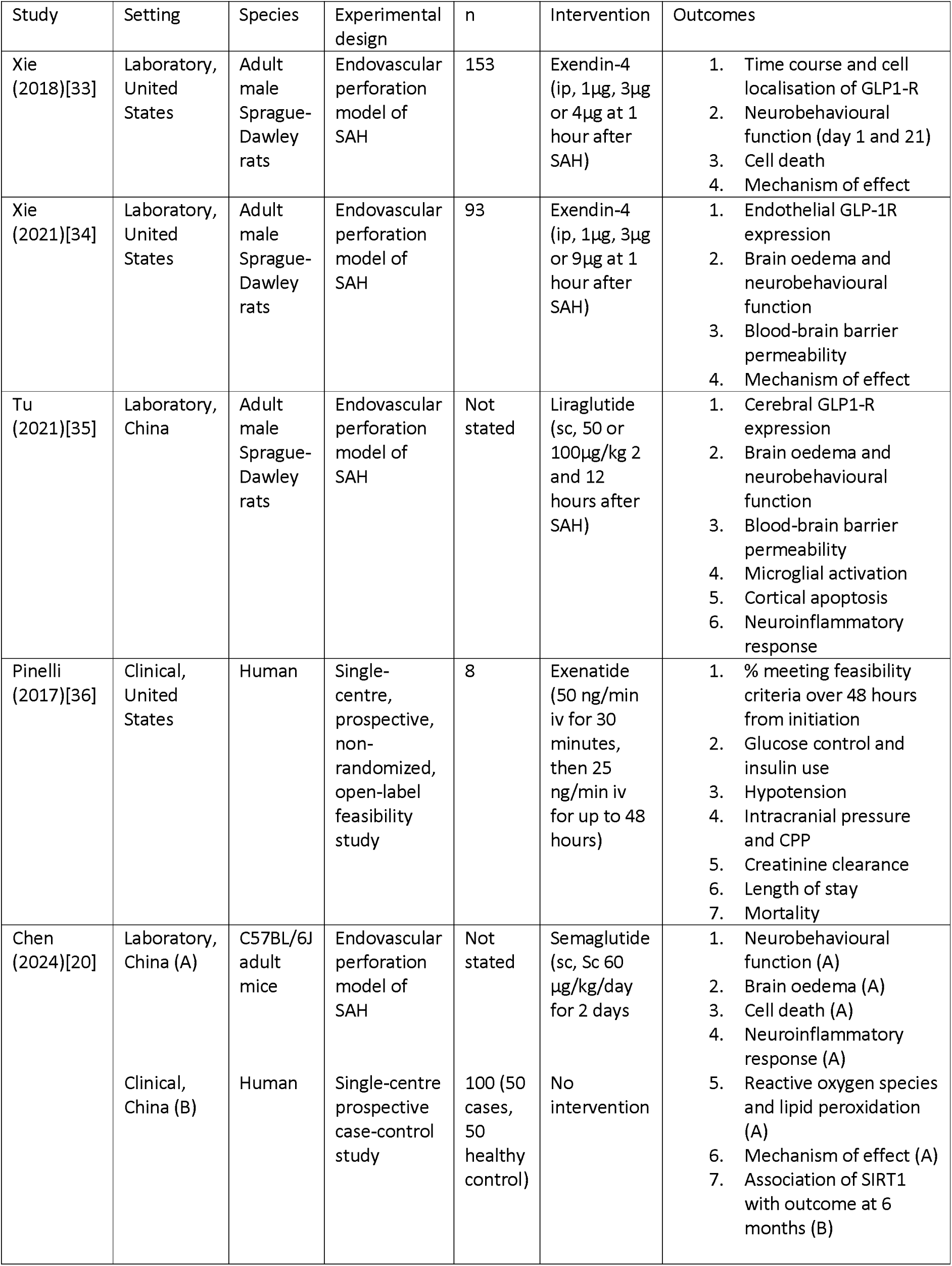
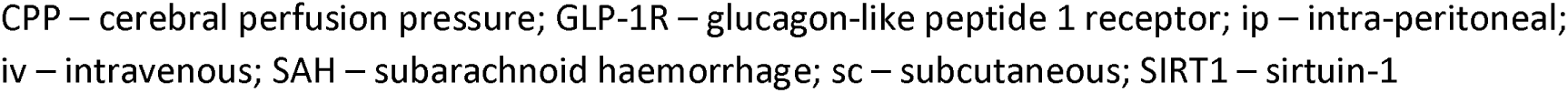
Key characteristics of included studies.

Review findings

Animal studies

### GLP-1 receptors (GLP-1R)

Xie at al. (2018) determined the time course of GLP-1 receptor expression after SAH, and localised GLP-1R at 24 hours post-ictus in rats. They demonstrated significantly elevated GLP-1R levels at three hours which peaked at 24 hours and had deceased at 72 hours. GLP-1R were expressed by neurons, astrocytes and microglia in ipsilateral basal cortex, with the greatest expression in neurons.^33^ Xie et al. (2021) also show elevation of GLP-1R expression at 24 hours (that was augmented by exendin-4) and further demonstrate expression of GLP-1R on endothelial cells of microvessels of the ipsilateral basal cortex.^34^ Xie et al. (2018) showed that 3μg of exendin-4 resulted in significant elevation of GLP-1R expression compared to sham and vehicle groups.^33^

Conversely, Tu et al. found that GLP-1R protein expression in rats was downregulated at 24 hours after SAH; however, they also found significant upregulation following GLP-1RA at a higher dose (100 μg.kg liraglutide, but not 50 μg.kg liraglutide).^35^

### Neurobehavioural function

All animal studies used the modified Garcia test to assess neurobehavioural function. Scores in six sub-domains (spontaneous activity, climbing ability, spontaneous movement of four limbs, forelimbs outstretching, vibrissa touch, and proprioception) determined by a blinded assessor are added together; higher scores reflect better function.

Xie et al. (2018), Xie et al. (2021) and Tu et al. showed that SAH reduced the score on the modified Garcia test compared with sham operated animals (rats).^33–35^ All (rats and mice) showed that GLP-1RA were associated with higher scores compared to vehicle at 24 hours after SAH; this was evident at 3μg and 9μg of exendin-4, the higher (100 μg.kg) liraglutide dose and 60μg.kg.day semaglutide.^20,33–35^

Xie et al. (2018) further found that SAH in rats (compared to sham) significantly reduced falling latency in rotarod tests at 7, 14 and 21 days. This was significantly improved by 3μg exendin-4 at all time points. Improved water maze results at day 21-25 were also shown.^33^

### Cerebral oedema

Three studies (two rat, one mouse) report on cerebral oedema, assessed using the wet-dry method to quantify brain water content.^20,34,35^ All showed a significant increase in brain water content 24-48 hours after SAH, compared to sham, which was reduced by treatment with each GLP-1RA at all doses investigated. In rats the cerebral oedema was restricted to the cerebral hemispheres, with the brainstem and cerebellum showing no increase in brain water content after SAH. ^34,35^

### Blood-brain barrier (BBB) permeability

Two studies in rats report on blood-brain barrier permeability.^34,35^ Both report an increase in Evans Blue dye extraction at 24-48 hours after SAH, indicating increased BBB permeability. Consistent with this Xie et al. (2021) also found increased expression of albumin and reduced expression of tight-junction related proteins after SAH.^34^ The SAH-induced increase in BBB permeability was significantly reduced by 3μg exendin-4 and both doses of liraglutide.

### Cell death

Two rat studies and one mouse study investigated cell death.^20,33,35^ Increased cell death was shown after SAH in all experiments, in ipsilateral cortex (rats) or hippocampus (mice). Cell death was significantly reduced by 3μg exendin-4 and both doses of liraglutide in rats, and by semaglutide in mice.

### Neuroinflammatory response

One study in rats and one in mice examine aspects of the post-SAH neuroinflammatory response.^20,35^ In mice, hippocampal levels of pro-inflammatory cytokines (TNFα, IL-6, NF-κB, IL-1β) were significantly increased by SAH; this increase was mitigated by semaglutide treatment.^20^ In rats, SAH-induced increases in COX-2, iNOS, TNFβ and IL-1β expression were attenuated by liraglutide at 100μg.kg.^35^

### Blood glucose

In rats, blood glucose was elevated after SAH. There was no significant difference in blood glucose between low, medium and high doses of exendin-4 in the first 24 hours.^33,34^

### Mechanisms of effect

Chen at al. investigated the role of ferroptosis (a mediator of cell death in SAH) and the SIRT1 pathway (a histone deacetylase that functions as a transcription regulator involved in the stress response) in mice. At 48 hours after SAH, SIRT1 levels were significantly reduced. Levels were increased by semaglutide. Knockout of SIRT1 by siRNA was associated with a diminished effect of semaglutide on brain water content and neurobehavioural scores. Further, semaglutide reduced reactive oxygen species and lipid peroxidation, and increased expression of proteins (FSP1 and GPX4) associated with ferroptosis, controlled by the Nrf2/HO-1 pathway. They conclude that semaglutide-mediated SIRT1 upregulation inhibited ferroptosis.^20^

Xie at al. (2018) examined the phosphatidylinositol 3-kinase (PI3K)/protein kinase B (Akt) apoptosis signalling pathway. They found SAH upregulates pro-apoptotic markers (Bax) and downregulates anti-apoptotic markers (Bcl-xl, Bcl-2) as well as PI3K and p-Akt. These changes were reversed by 3μg exendin-4 administration, an effect that was blocked by GLP-1R knockdown with siRNA. The effects of exendin-4 were also inhibited by pre-treatment with LY294002, a non-specific PI3K inhibitor. They conclude that the PI3K/Akt pathway is involved in the neuroprotective effect of exendin-4.^33^

Xie at al. (2021) show that GLP-1R siRNA inhibits the upregulation of GLP-1R in response to exendin-4 and reverses the benefits of exendin-4 on neurobehavioural function. They also demonstrate that SAH-induced expression of AMPK was increased by exendin-4, an effect that is GLP-1R dependent. Similarly, SAH-induced changes in NF-κB and MMP-9 were reversed by GLP-1R dependent effects of exendin-4. Finally, the preservation of BBB tight-junction-related protein expression (claudin-5 and occludin) seen with exendin-4 treatment was abolished by pre-treatment with GLP1-R siRNA. They conclude that GLP1-R/AMPK-dependent NF-κB/MMP9 inhibition is a potential neuroprotective mechanism after SAH.^34^

### Human studies Interventional

Pinelli and colleagues conducted a single centre prospective study to test the feasibility of exenatide infusion for the management of hyperglycaemia after acute brain injury.^36^ Of the eight adult patients recruited one had aSAH. During the 48-hour intravenous infusion of exenatide 87.5% of patients achieved the blood glucose goal, with none experiencing severe hypoglycaemia or nausea requiring discontinuation. There were four episodes of hypoglycaemia in three patients and no other safety concerns were identified. All-cause mortality was 37.5%. The trial ended early with no reason given for this. Results for the patient with aSAH are not reported separately.^37^

### Observational

Chen et al. compared SIRT1 levels in fifty aSAH patients and 50 healthy controls.^20^ Levels in controls were higher than in good Hunt and Hess grade aSAH patients, and both were higher than levels in poor grade aSAH patients. Patients with good outcomes (scores of 3-6 on the modified Rankin scale, mRS) had higher SIRT1 levels than those with poor (mRS 0-2) outcomes. They conclude that SIRT1 levels at 48 hours post-ictus were associated with clinical severity (Hunt and Hess) and 6 month outcome (dichotomised mRS) and could be used to predict outcome. Exposure to GLP-1 receptor agonists in the cohort is not reported.

## Discussion

A limited number of animal studies in rodents using the endovascular perforation model of aneurysmal subarachnoid haemorrhage are consistent in showing benefit of GLP-1RA on neurobehavioural function, blood-brain barrier permeability and cerebral oedema, neuroinflammation and cell death. These effects are shown to be GLP-1R dependent and to involve signalling pathways likely to be associated with early brain injury in aSAH.^38–42^ These effects are present in two species and with three drugs and dose schedules, with evidence of a dose-response effect for some drugs and outcomes.

There are no human studies of GLP-1RA for neuroprotection after aSAH. There is evidence of an association between SIRT1 and outcome in humans. This is a pathway considered relevant to early brain injury and a finding that replicates one animal study, but it cannot be concluded from this that GLP-1RA would show benefits in clinical practice. A single small study of short term exenatide infusion demonstrates that short term intravenous infusions are feasible and safe in critically ill adults with acute brain injury, but reports no data for the one SAH patient, or for patient-centred outcomes.

Taken together the studies demonstrate that there is potential for GLP-1RA to be neuroprotective after SAH, but that whether will ultimately translate into benefits for patients cannot currently be assessed. Nor is it possible to determine the ideal drug and dose regimen, although the consistency of effect and demonstration of GLP-1R mediated mechanisms, suggests that this is likely to be a class effect. In this our findings mirror those of reviews of GLP-1RA for neuroprotection in other forms of acute brain injury, in terms of species investigated, potential for benefit, broad mechanisms (such as reduction in apoptosis or cerebral oedema) and some specific pathways (PI3k/Akt for example).^43,44^ We also note that drugs in other classes whose action is medicated via GLP-1R show neuroprotective effects in early brain injury after aSAH.^45^ However, as a central finding of this review is that the evidence base is limited, considerable work is required before definitive studies can be undertaken.

### Limitations of the review

The particular animal model (endovascular perforation) is commonly used although is not entirely standardised, with variations in surgical technique (that may affect cerebral blood flow) and the device used for perforation. As with clinical aSAH the perforation occurs at mean arterial pressure, and results in variable blood load and distribution, although always within the anterior portion of the Circle of Willis. However this model does not incorporate aneurysm formation and whether this alters pathophysiology and outcomes is an open question. Further, the development of delayed cerebral ischaemia in this model is unpredictable; while this may mirror the clinical situation it makes study of this crucial component of disease difficult.^46^

It may then be that slight differences in perforation technique, or in laboratory methods, underlie the divergence of findings on GLP-1R expression after SAH in rats.^33,35^ Further clarification of expression both in time and space would be advisable, ideally in human health and disease as well. Other results from animal studies are consistent, and we do not consider this one point of difference to undermine the overall conclusion.

It is also the case that rodents are different species to humans. Alternative large animal and non-human primate models are available but are not in widespread use and are subject to the same caveats as rodent models. In addition to those above, controlled laboratory conditions in otherwise healthy animals do not match the clinical presentation of aneurysmal subarachnoid haemorrhage in adults who frequently have co-morbidities. It remains a concern that inadequate models may be contributing to the lack of successful translation of apparently promising neuroprotective therapies.^47,48^

Drug administration post-ictus better replicates the clinical setting, although the timing (1-2 hours after SAH) is difficult to match in many if not all health care systems. In keeping with the experimental focus on early brain injury and the available animal models the drug regimens are short term and it remains unknown whether more (or less) benefit would be derived with longer courses of treatment.

With the exception of Xie et al. (2018) who studied neurobehavioural function at 3-4 weeks after onset, there is no animal data beyond 48 hours.^33^ This is important for aSAH, as the period of risk for brain injury extends well beyond the initial insult. Clinically this is typically considered in two phases: early brain injury (EBI, first three days) and delayed neurological deterioration including deficits related to delayed cerebral ischaemia (DCI) for the first three weeks.^2,15^ Unfortunately, most preclinical models (including rodent) do not replicate DCI well.^48,49^ The findings of Xie et al. (2018) of benefit maintained at the later time point could reflect a reduction in EBI alone, or a reduction in both EBI and DCI.^33^ In the absence of imaging and other techniques to identify ischaemia, vasospasm and spreading depolarisations the effect of GLP-1RA on these different components of brain injury after SAH cannot be determined, although finding that exendin-4 preserves cerebral microcirculation after ischaemic stroke offers hope for an effect on DCI.^50^

We have identified a lack of data on use of GLP-1RA in patients with aSAH, whether pre-or post-ictus. There is evidence to support a protective effect of pre-ictus GLP-1RA use on the incidence of stroke more generally, although this may only be relevant for patients with diabetes, and may not hold for aSAH specifically.^51–53^ The effect of GLP-1RA use for neuroprotection after acute ischaemic stroke is under investigation, but we were unable to identify studies in aSAH.^54,55^

Implications of the findings for research

1. Animal

We consider the following to be priorities for pre-clinical research:

a. Longer term neurobehavioural studies to better define functional outcomes.
b. Assessment of the effect on delayed cerebral ischaemia as well as early brain injury in appropriate models.
c. Comparison of drugs and dosing regimens to determine the ideal intervention.

2. Human

To establish the potential for benefit in clinical studies we suggest:

a. Epidemiological studies of the effect of GLP-1RA exposure on the incidence of and outcomes from aSAH.
b. Basic science studies of underlying mechanisms of action identified in pre-clinical studies.
c. Phase 2 studies demonstrating feasibility of administration and effects on biomarkers of disease as surrogate outcomes.

We do not recommend phase 3 studies without additional supporting evidence, but with the known inadequacies of animal models noted above would suggest that research in humans is prioritised.

### Implications of the findings for practice

Extensive further research is needed before GLP-1RA can be considered for use for neuroprotection after aneurysmal subarachnoid haemorrhage in clinical practice.

## Conclusions

A very limited amount of animal evidence using a rodent endovascular perforation model suggests GLP-1RAs may be neuroprotective after aSAH. There is a lack of evidence from large animals or humans to either confirm or refute this. Further research is justified given the lack of available neuroprotective treatments for aneurysmal subarachnoid haemorrhage and the safety in real world use of GLP-1RAs.^56^

## Abbreviations

Akt: Activation of protein kinase B
AMPK: AMP-activated protein kinase
aSAH: Aneurysmal subarachnoid haemorrhage
Bax: Bcl-2-like protein 4
BBB: Blood brain barrier
Bcl-2: B-cell lymphoma 2 protein
Bcl-xl: B-cell lymphoma extra large protein
COX-2: Cyclo-oxygenase 2
DCI: Delayed cerebral ischaemia
EBI: Early brain injury
FSP1: Ferroptosis suppressor protein 1
GLP-1: Glucagon-like peptide-1
GLP-1R: Glucagon-like peptide-1 receptor
GLP-1RA: Glucagon-like peptide-1 receptor agonist
GPX4: Glutathione peroxidase 4
HO-1: Haem oxygenase 1
IL-1β: Interleukin 1-beta
IL-6: Interleukin 6
ICTRP: International Clinical Trials Registry Platform
iNOS: Inducible nitric oxide synthase
JBI: Joanna Briggs Institute
MMP9: Matrix metalloproteinase 9
mRS: Modified Rankin Scale
NF-κB: Nuclear factor kappa-light-chain-enhancer of activated B cells
Nrf2: Nuclear factor erythroid 2-related factor 2
PI3K: Phosphoinositide 3-kinase
PRISMA: Preferred reporting items for systematic review and meta-analyses
PROSPERO: Prospective register of systematic reviews
siRNA: Small interfering ribonucleic acid
SIRT1: Sirtuin 1
TNFα: Tumour necrosis factor alpha
TNFβ: Tumour necrosis factor beta

## Data Availability

All data included in the present study are available in the references papers and summarised in the manuscript.

## Acknowledgements

No acknowledgements.

## Author contributions

See CreDIT statement.

## Availability of data, code, and other materials

All relevant data, code and other materials is provided in the manuscript, appendices and supplementary material.

## Appendices

### Appendix I: Search strategy

1. Embase (Ovid)

Embase <1974 to 2025 June 11>

1. · subarachnoid hemorrhage/ 59010
2. · exp brain hemorrhage/ 202241
3. · exp carotid artery aneurysm/ 6604
4. · exp brain hematoma/ 34574
5. · ((subarachnoid* or arachnoid* or brain* or cerebral or intracerebral or intracranial or cerebell* or intraventricular or parenchymal or infratentorial or supratentorial or carotid) adj4 (h?emorrhag* or h?ematoma or bleed* or blood or vasospasm* or vasoconstrict* or angiospasm*)).ti,ab,kf. 317784
6. · exp brain vasospasm/ 10502
7. · “delayed cerebral isch?emia”.ti,ab,kf. 2992
8. · ((cortical or cortex or spreading) adj3 (depolarisation or depression)).ti,ab,kf. 4574
9. · spreading cortical depression/ 3508
10. · 1 or 2 or 3 or 4 or 5 or 6 or 7 or 8 or 9 427939
11. · (exenatide or “AC 2993” or AC2993 or “ITCA 650” or ITCA650 or “exendin 4”).ti,ab,kf. 8149
12. · exendin 4/ 13765
13. · exenatide.dy. 0
14. · exendin 4.dy. 13772
15. · liraglutide/ 15846
16. · (liraglutide or “NN 2211” or NN2211 or “NNC 90 1170” or “NNC90 1170”).ti,ab,kf. 9299
17. · liraglutide.dy. 16112
18. · albiglutide/ 1695
19. · (albiglutide or GSK 716155 or GSK716155 or (albumin adj3 (“GLP 1” or GLP1 or “glucagon like peptide”))).ti,ab,kf. 529
20. · albiglutide.dy. 1695
21. · dulaglutide/ 4057
22. · (dulaglutide or LY2189265 or “LY 2189265”).ti,ab,kf. 2033
23. · dulaglutide.dy. 4057
24. · lixisenatide/ 2627
25. · (lixisenatide or “AVE 0010” or AVE0010).ti,ab,kf. 1162
26. · lixisenatide.dy. 2849
27. · semaglutide.dy. 8214
28. · semaglutide/ 8214
29. · (semaglutide or “NN 9535” or NN9535).ti,ab,kf. 5294
30. · tirzepatide/ 2253
31. · Tirzepatide.dy. 2253
32. · (Tirzepatide or “LY-3298176” or LY3298176 or Mounjaro).ti,ab,kf. 1672
33. · ((glucagon like peptide* or “GLP 1” or GLP1) adj3 (analog* or agonist*)).ti,ab,kf. 21919
34. · “Incretin mimeti*”.ti,ab,kf. 777
35. · (“GLP1-RA” or GLP1RA or “GLP-1-RA” or “GLP-1” or “GLP-1R” or “GLP-1RA”).ti,ab,kf. 34788
36. · glucagon like peptide 1 receptor agonist/ 16459
37. · or/11-36 61030
38. · 10 and 37 532

2.Medline All (Ovid)

Ovid MEDLINE(R) ALL <1946 to June 12, 2025>

1. · exp Subarachnoid Hemorrhage/ 25474
2. · exp Intracranial Hemorrhages/ 85049
3. · Intracranial Aneurysm/ 33717
4. · exp Hematoma, Subdural/ 10523
5. · ((subarachnoid* or arachnoid* or brain* or cerebral or intracerebral or intracranial or cerebell* or intraventricular or parenchymal or infratentorial or supratentorial or carotid) adj4 (h?errhag* or h?ematoma or bleed* or blood or vasospasm* or vasoconstrict* or angiospasm*)).ti,ab,kf. 152919
6. · Vasospasm, Intracranial/ 4042
7. · “delayed cerebral isch?emia”.ti,ab,kf. 2061
8. · ((cortical or cortex or spreading) adj3 (depolarisation or depression)).ti,ab,kf. 3565
9. · Cortical Spreading Depression/ 2320
10. · 1 or 2 or 3 or 4 or 5 or 6 or 7 or 8 or 9 249688
11. · (exenatide or “AC 2993” or AC2993 or “ITCA 650” or ITCA650 or “exendin 4”).ti,ab,kf. 4496
12. · Exenatide/ 3083
13. · Liraglutide/ 2944
14. · (liraglutide or “NN 2211” or NN2211 or “NNC 90 1170” or “NNC90 1170”).ti,ab,kf. 4731
15. · (albiglutide or GSK 716155 or GSK716155 or (albumin adj3 (“GLP 1” or GLP1 or “glucagon like peptide”))).ti,ab,kf. 282
16. · (dulaglutide or LY2189265 or “LY 2189265”).ti,ab,kf. 988
17. · (lixisenatide or “AVE 0010” or AVE0010).ti,ab,kf. 646
18. · (semaglutide or “NN 9535” or NN9535).ti,ab,kf. 2951
19. · (Tirzepatide or “LY-3298176” or LY3298176 or Mounjaro).ti,ab,kf. 1030
20. · ((glucagon like peptide* or “GLP 1” or GLP1) adj3 (analog* or agonist*)).ti,ab,kf. 13414
21. · “Incretin mimeti*”.ti,ab,kf. 472
22. · (“GLP1-RA” or GLP1RA or “GLP-1-RA” or “GLP-1” or “GLP-1R” or “GLP-1RA”).ti,ab,kf. 20533
23. · Glucagon-Like Peptide-1 Receptor Agonists/ 3563
24. · 11 or 12 or 13 or 14 or 15 or 16 or 17 or 18 or 19 or 20 or 21 or 22 or 23 28714
25. · 10 and 24 205

3. Cochrane CENTRAL (Wiley)

ID Search Hits

#1 MeSH descriptor: [Subarachnoid Hemorrhage] explode all trees 874 #2 MeSH descriptor: [Intracranial Hemorrhages] explode all trees 3251 #3 MeSH descriptor: [Intracranial Aneurysm] explode all trees 695

#4 MeSH descriptor: [Hematoma, Subdural] explode all trees 238

#5 (((subarachnoid* or arachnoid* or brain* or cerebral or intracerebral or intracranial or cerebell* or intraventricular or parenchymal or infratentorial or supratentorial or carotid)

Near/4 (h?errhag* or h?ematoma or bleed* or blood or vasospasm* or vasoconstrict* or angiospasm*))):ti,ab,kw 16986

#6 MeSH descriptor: [Vasospasm, Intracranial] explode all trees 227 #7 “delayed cerebral” NEXT isch?emia 272

#8 (((cortical or cortex or spreading) NEAR/3 (depolarisation or depression))):ti,ab,kw 451 #9 MeSH descriptor: [Cortical Spreading Depression] explode all trees 12

#10 #1 or #2 or #3 or #4 or #5 or #6 or #7 or #8 or #9 19892

#11 ((exenatide or “AC 2993” or AC2993 or “ITCA 650” or ITCA650 or “exendin 4”)):ti,ab,kw 1417

#12 MeSH descriptor: [Exenatide] explode all trees 681 #13 MeSH descriptor: [Liraglutide] explode all trees 1018

#14 ((liraglutide or “NN 2211” or NN2211 or “NNC 90 1170” or “NNC90 1170”)):ti,ab,kw 2506

#15 ((albiglutide or “GSK 716155” or GSK716155 or (albumin NEAR/3 (“GLP 1” or GLP1 or “glucagon like peptide”)))):ti,ab,kw 155

#16 ((dulaglutide or LY2189265 or “LY 2189265”)):ti,ab,kw 595 #17 ((lixisenatide or “AVE 0010” or AVE0010)):ti,ab,kw 384

#18 ((semaglutide or “NN 9535” or NN9535)):ti,ab,kw 1590

#19 ((Tirzepatide or “LY-3298176” or LY3298176 or Mounjaro)):ti,ab,kw 478

#20 (((glucagon like peptide* or “GLP 1” or GLP1) NEAR/3 (analog* or agonist*))):ti,ab,kw 4079

#21 (Incretin NEXT mimeti*):ti,ab,kw 45

#22 ((“GLP1-RA” or GLP1RA or “GLP-1-RA” or “GLP-1” or “GLP-1R” or “GLP-1RA”)):ti,ab,kw 5240

#23 MeSH descriptor: [Glucagon-Like Peptide-1 Receptor Agonists] explode all trees 316

#24 #11 or #12 or #13 or #14 or #15 or #16 or #17 or #18 or #19 or #20 or #21 or #22 or #23 10288

#25 #10 and #24 82

4. WHO ICTRP

1. (hemorrhag* or haemorrhage or hematom* or haematom*) AND (exenatide or liraglutide or albiglutide or dulagutide or lixisenatide or semaglutide or tirzepatide) 0
2. (exenatide or liraglutide or albiglutide or dulagutide or lixisenatide or semaglutide or tirzepatide) and (subarachnoid* or arachnoid* or brain* or cerebral or intracerebral or intracranial or cerebell* or intraventricular or parenchymal or infratentorial or supratentorial or carotid) 44
3. (“glucagon like peptide*” or “GLP 1” or GLP1) and (subarachnoid* or arachnoid* or brain* or cerebral or intracerebral or intracranial or cerebell* or intraventricular or parenchymal or infratentorial or supratentorial or carotid) 24

### Appendix II: Data extraction instrument

Paper for inclusion reference number Evidence source details/characteristics Citation

Species Setting

Clinical / laboratory Type of evidence source

Study design/characteristics Design

Participants (numbers, age, sex etc) Intervention

Comparator Outcome(s) Results extracted

Outcome (describe) Data

Outcome (describe) Data

Outcome (describe) Data

Outcome (describe) Data

Outcome (describe) Data

## CRediT Author Contribution Statement

Aidan Marsh Data curation, Investigation, Writing – review & editing

Alex Mortimer Conceptualization, Writing – review & editing

Aravind Ramesh Writing – review & editing

Lucy Holmes Writing – review & editing

Mario Teo Writing – review & editing

Matt Thomas Conceptualization, Data curation, Investigation, Writing – original draft, Writing – review & editing

Ruth Cookson Writing – review & editing

Svetlana Mastitskaya Conceptualization, Writing – original draft, Writing – review & editing

Sarah Rudd Resources, Writing – original draft

Sam Parker Data curation, Investigation, Writing – original draft, Writing – review & editing

Preferred Reporting Items for Systematic reviews and Meta-Analyses extension for Scoping Reviews (PRISMA-ScR) Checklist

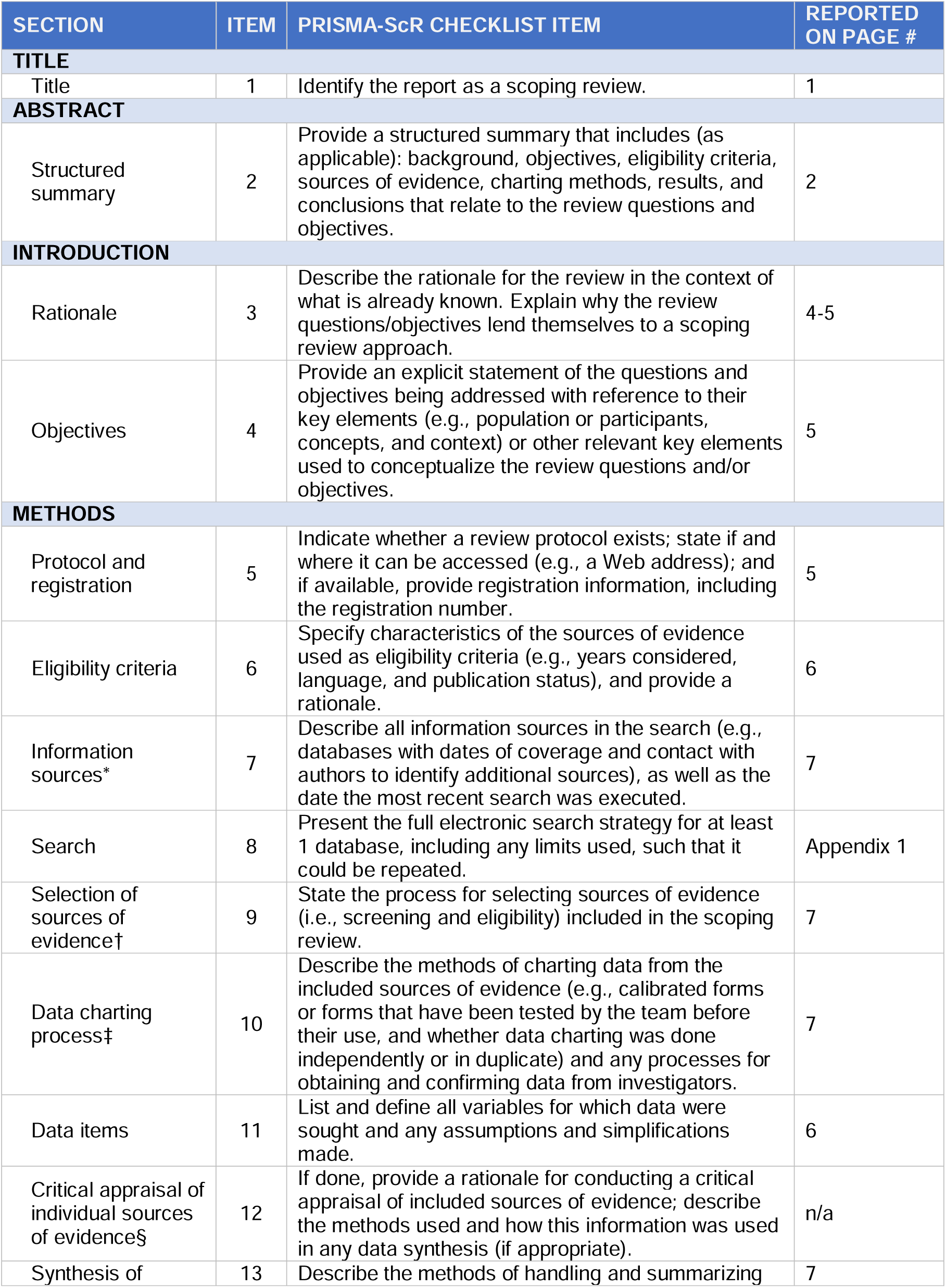
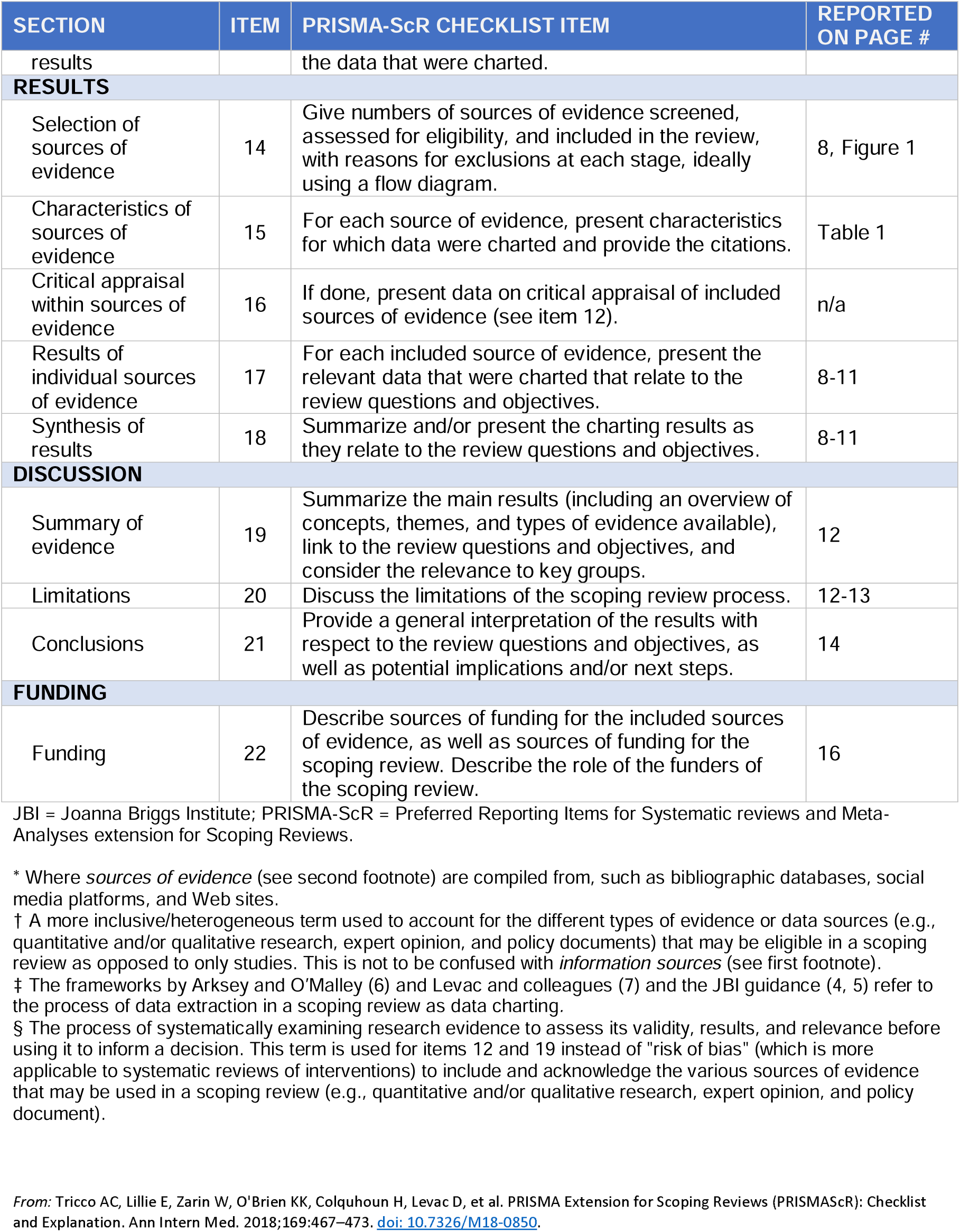

